# PROCC: a predictive score to identify KRAS wild type metastatic colorectal cancer patients who are likely to benefit from panitumumab treatment

**DOI:** 10.1101/2024.12.18.24319211

**Authors:** CM Galmarini, R Zamora, P Gómez del Campo, J Castillo Izquierdo, JA De All, JM Domínguez

## Abstract

**Background:** Practice guidelines recommend using panitumumab in combination with chemotherapy to treat KRAS wild-type (WT) metastatic colorectal cancer (mCRC) patients where it was shown to significantly extend progression-free survival (PFS) and overall survival (OS). Still, a proportion of patients will not achieve this goal. We propose a simplified predictive score to identify patients who are likely to benefit from panitumumab treatment.

**Methods:** NCT00364013 (TRDS) (n=460) was used as training dataset and NCT00339183 (VALDS) (n=479) as validation set. Datasets were obtained from www.projectdatasphere.org and included KRAS WT mCRC patients treated with panitumumab in combination (P/FOL) or not with FOLFOX (FOL) (TRDS) or FOLFIRI (VALDS) as 1st and 2nd line therapy. TRDS was used to generate synthetic representations (SRs) for each patient through the integration of 36 clinical and analytical features collected, respectively, during the screening phase and the first month of inclusion. These SRs were then input into a machine learning (ML) framework to identify subgroups of patients based on their similarities. The resultant subpopulations were correlated with PFS and OS. Differential variables between subgroups were identified through feature contribution analysis and included in a multivariable logistic regression model. Independent predictive factors found to be statistically significant were used to generate a predictive score of panitumumab response at baseline that was validated in VALDS.

**Results:** ML identified two different subpopulations on the TRDS: SPA (n=162) and SPB (n=298). Only SPA patients had a lower risk of death when treated with P/FOL compared to FOL (HR 0.68 95%CI 0.48-0.99; p=.04). Patients in SPB showed no significant differences on OS between P/FOL and FOL (p=.27). Feature contribution analysis identified 15 differential features between both subpopulations. From these, CEA, ALP, LDH, and platelets were selected to create a simplified predictive score for panitumumab response ranging 0-18. When applied to TRDS, this score yielded an area under the curve of 0.81 (95% CI: 0.77 to 0.85). A score ≥8.5 was correlated to a lower risk of progression (HR 0.67 95% CI 0.47-0.97; p=.03) and death (HR 0.65 95%CI 0.43-0.98; p=.04) after P/FOL compared to FOL. No significant differences were observed for PFS and OS between P/FOL and FOL in patients with a score <8.5. The predictive score was then validated in the VALDS set with similar results (score ≥8.5: PFS: HR 0.48 95%CI 0.33-0.70; p=.002; OS: HR 0.60 95%CI 0.42-0.87; p=.007; score <8.5, PFS: p=.2; OS: p=.9).

**Conclusions:** Based on CEA, ALP, LDH and platelet baseline levels, this easily applicable predictive score might be helpful to accurately select KRAS WT mCRC patients who would benefit from addition of panitumumab to chemotherapy treatment in first- or second-line therapy. Further work is required to validate this approach in prospective cohorts of patients.

## INTRODUCTION

The therapeutic landscape for stage IV metastatic colorectal cancer (mCRC) has evolved significantly over the past decades, offering new hope for improving patient outcomes^1^. Standard first-line treatments have historically been based on chemotherapy combinations, such as 5-fluorouracil (5-FU)/leucovorin (LV) with oxaliplatin (FOLFOX) or irinotecan (FOLFIRI)^2–4^. More recently, these regimens have been enhanced by integrating targeted therapies, including multikinase inhibitors (e.g., regorafenib), B-Raf inhibitors (e.g., encorafenib), antiangiogenic agents (e.g., bevacizumab), anti-epidermal growth factor receptor (EGFR) monoclonal antibodies (e.g., cetuximab, panitumumab), and immune checkpoint inhibitors (e.g., pembrolizumab, nivolumab)^5, 6^. These advancements underscore the growing importance of molecular testing, which guides therapeutic decisions by identifying key genetic mutations (e.g., RAS, BRAF, MSI) and tailoring treatments accordingly^7^. This shift toward targeted therapies continues to improve progression-free survival (PFS) and overall survival (OS), which now range between 9-12 months and 28-32 months for PFS and OS, respectively, in different studies^8, 9^. Therefore, the identification of novel biomarkers offers opportunities to better select patients for targeted treatments, addressing primary resistance mechanisms and improving outcomes.

For patients with KRAS wild-type (WT) mCRC, treatment with anti-EGFR antibodies combined with cytotoxic chemotherapy (FOLFOX or FOLFIRI) or as monotherapy remains a cornerstone of therapy^10^. Among these agents, panitumumab, a fully human monoclonal antibody, has proven particularly beneficial in this clinical setting. Several clinical studies have demonstrated that adding panitumumab to chemotherapy significantly improves overall response rates (ORR) and survival outcomes in some cases^11–13^. Different meta-analyses have further validated the survival advantages of panitumumab when combined with chemotherapy^14–16^. Despite the importance of KRAS status in guiding the use of panitumumab, it does not definitively predict whether an individual patient will benefit from this therapy. Determining which patients should receive this anti-EGFR agent, either as monotherapy or in combination with chemotherapy, remains a subject of ongoing debate and research. Panitumumab’s efficacy varies across treatment lines and tumor locations. While first-line therapy with panitumumab shows response rates around 45%-50%, its efficacy drops to 15%-25% in second line and 10%-15% in third-line settings^12, 17, 18^. Moreover, panitumumab is more effective in left-sided tumors, whereas right-sided tumors exhibit limited response, adding further challenges in selecting the appropriate patient population for this treatment^19, 20^. This discrepancy in response, combined with toxicity risks such as skin rash, paronychia, fatigue, nausea and/or diarrhea, emphasizes the need for better patient selection, potentially through novel biomarkers, to maximize benefit and minimize unnecessary adverse effects.

To address this gap, we developed and evaluated PROCC (Panitumumab Response Optimizer for Colorectal Cancer patients), an accessible, sensitive predictive tool that utilizes four common laboratory metrics to forecast the likelihood of benefit from panitumumab treatment in KRAS WT mCRC. We first applied machine learning (ML) techniques to uncover patterns and features associated with patient clinical outcome, which were then used as input variables to build the predictive score. The score was then validated using a large cohort of patients from two distinct clinical trials involving KRAS WT mCRC patients. Our findings suggest that PROCC accurately identifies, before treatment, patients who are likely to benefit from panitumumab therapies.

## MATERIAL AND METHODS

### Patient population

The patient population included KRAS WT mCRC patients treated or not with panitumumab in combination with FOLFOX4 (NCT00364013; PRIME) or FOLFIRI (NCT00339183) as first- or second-line therapy clinical trials^11, 12^. Datasets were obtained from www.projectdatasphere.org. The NCT00364013 trial determined the treatment effect of panitumumab in combination with FOLFOX4 compared to FOLFOX4 alone as first line therapy for mCRC. The NCT00339183 evaluated the treatment effect of panitumumab plus FOLFIRI compared to FOLFIRI alone as second line therapy for mCRC. More information on the trials can be found at www.clinicaltrials.gov. In both trials, datasets consisted of a randomly selected 80% sample of the overall patient population. From those, we have selected only those that were confirmed as KRAS WT. NCT00364013 (TRDS) (n=460) was used as training dataset while NCT00339183 (VALDS) (n=479) was used as validation set.

### Input data on TRDS (NCT00364013 dataset)

The following variables were available for all individuals: sex, age in years at screening, weight (Kg), height (cm), body surface area (BSA; m^2^), body mass index (BMI), race, ECOG performance status, primary tumor diagnosis, histological subtype, actual treatment, prior surgery, metastases to liver at study entry, adverse events type, adverse event severity (in grade), number of days on toxicity, albumin (g/L), alkaline phosphatase (ALP; UI/L), creatinine (µmol/L), lactate dehydrogenase (LDH; UI/L), hemoglobin (g/L), platelets (10^9^/L), white blood cells (WBC; 10^9^/L), carcinoembrionary antigen (CEA; µg/L), lesion category (target vs. non-target), type of lesion (new vs. previous), sum of long diameter within lesion (mm), longest diameter (mm), theoretical panitumumab dose (mg/kg/cycle 1), theoretical oxaliplatinum dose (mg/kg/cycle 1), theoretical 5-FU dose (mg/kg/cycle 1), theoretical LV dose (mg/kg/cycle 1), theoretical panitumumab dose intensity, theoretical oxaliplatin dose intensity, theoretical 5-FU dose intensity and theoretical LV dose intensity. Adverse events (AEs) were graded using the Common Terminology Criteria for Adverse Events (version 3.0). Dose intensity was calculated as total dose per patient/(total days under treatment/7). All these variables were collected during the screening phase and the first month of inclusion on the trial.

### ML Model Development

Synthetic representations (SRs) for each patient were generated through the integration of the 36 variables on a feature vector. These SRs were then used as input vectors into a non-supervised ML framework. To extract nonlinear relationships of the data, we used unsupervised learning and UMAP, a dimensionality reduction technique, from the umap-learn package version 0.5.3, to place the SRs embeddings in a three-dimensional space. The space was then clustered using the DBSCAN algorithm from the scikit-learn package version 1.2.2. The Python version used was 3.10.9. The resultant subpopulations were correlated with OS. Differential variables between subgroups were identified through feature contribution analysis. Independent predictive factors found to be statistically significant were used to generate a predictive score of panitumumab response at baseline that was validated in the test sets.

### Predictive score development

A multivariable logistic regression analysis was conducted to identify variables significantly associated with the highest benefit from panitumumab. Baseline values of variables with statistical significance (p<.05) in the feature contribution analysis were incorporated into the logistic regression model, employing a backward stepwise approach to derive the final predictors. The independent predictive factors that retained statistical significance in the final logistic regression analysis were utilized to compute the predictive score. Model performance was assessed using the area under the receiver operating characteristic (ROC) curve to evaluate discrimination, and the Hosmer-Lemeshow test to examine goodness-of-fit and calibration. The prediction of the subgroup benefiting from panitumumab was based on a prognostic score derived from odds ratios (OR) obtained through logistic regression analysis. The optimal cut-off value for the score was determined using Youden’s index, complemented by an evaluation of the accuracy at that threshold to ensure robust classification.

### Statistical analysis

All statistical analyses were performed using R version 4.3.2 and the corresponding packages therein as described below. Comparisons between categorical variables were based on the chi-square test while comparisons between continuous variables were based on the Mann-Whitney U test. Tests were performed with stats base package. PFS and OS were calculated from the date of randomization to the date of progression or death from any cause, respectively, or to the last follow-up examination (censored event). PFS and OS curves were calculated according to the Kaplan-Meier method and compared using the log-rank test. Survival analysis was performed using R survival package version 3.5-7. Unless otherwise explained, differences were considered significant if the p-value was <.05 (two-sided).

## RESULTS

### Development of an unsupervised ML model

We initially developed a ML model using SRs, which depicted data from 460 patients in the TRDS. The baseline characteristics of this patient cohort are shown in Supplementary Table 1 and Supplementary Figure 1. The ML model stratified patients into two distinct subpopulations according to the 36 clinical and analytical features integrated in the SRs: subpopulation A (SPA; N=162) and subpopulation B (SPB; N=298) (Figure 1). In SPA, 79 patients received panitumumab/FOLFOX4, while 83 were treated with FOLFOX4 alone. SPB comprised 153 patients treated with panitumumab/FOLFOX4 and 145 patients with FOLFOX4 alone.

**Figure 1.**
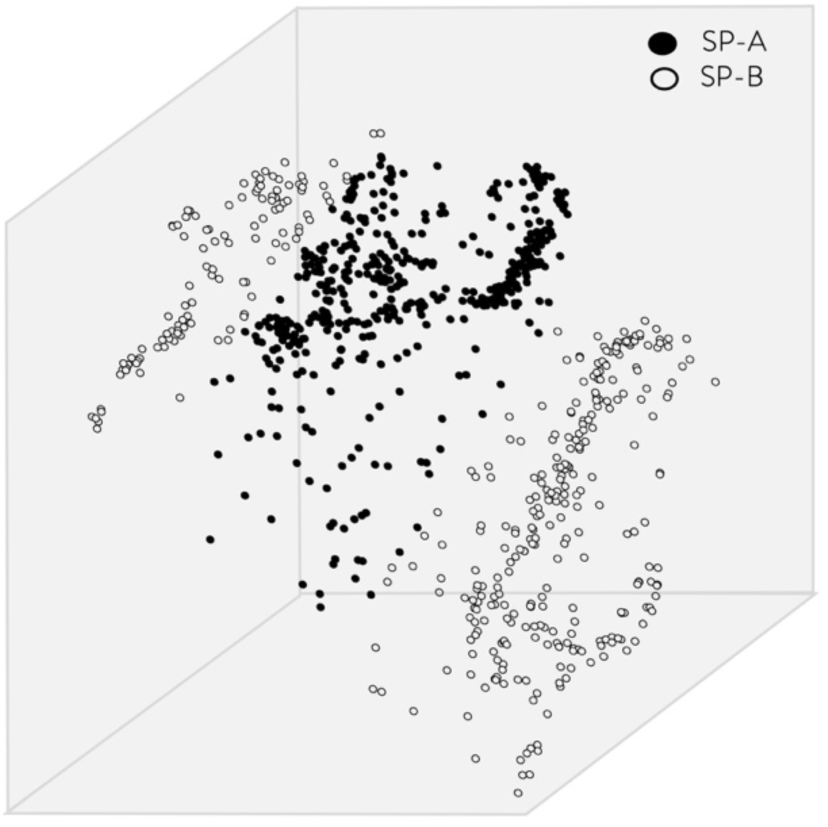
Embedding space and clustering of patients belonging to the training dataset (TRDS) by a machine learning framework. Each dot in the graph corresponds to the synthetic representation (SR) of each patient of the dataset. The SR was obtained through the integration of 36 clinical and analytical features collected, respectively, during the screening phase and the first month of inclusion on the trial. Two major clusters were identified: subpopulation A (SPA; n=162) and subpopulation B (SPB; n=298).

Subsequently, survival analyses were conducted for both subpopulations based on treatment with panitumumab/FOLFOX4 or FOLFOX4 alone. As shown in Figure 2A, SPA patients treated with panitumumab/FOLFOX4 exhibited a significantly reduced risk of death (HR 0.68; 95% CI 0.48-0.99; p=.04), with a median OS of 23.6 months (95% CI 18.1-29.8), compared to those receiving FOLFOX4 alone (median OS 17.1 months; 95% CI 13.6-19.0). Conversely, SPB patients showed no significant difference in OS between the two treatments (HR 0.92; 95% CI 0.69-1.23; p=.5), with median OS of 27.1 months (95% CI 18.9-29.5) for panitumumab/FOLFOX4 and 24.1 months (95% CI 19.1-30.2) for FOLFOX4 (Figure 2B). No significant differences were observed in PFS between patients treated with panitumumab/FOLFOX4 and those treated with FOLFOX4 alone within either subpopulation (Supplementary Figure 2).

**Figure 2.**
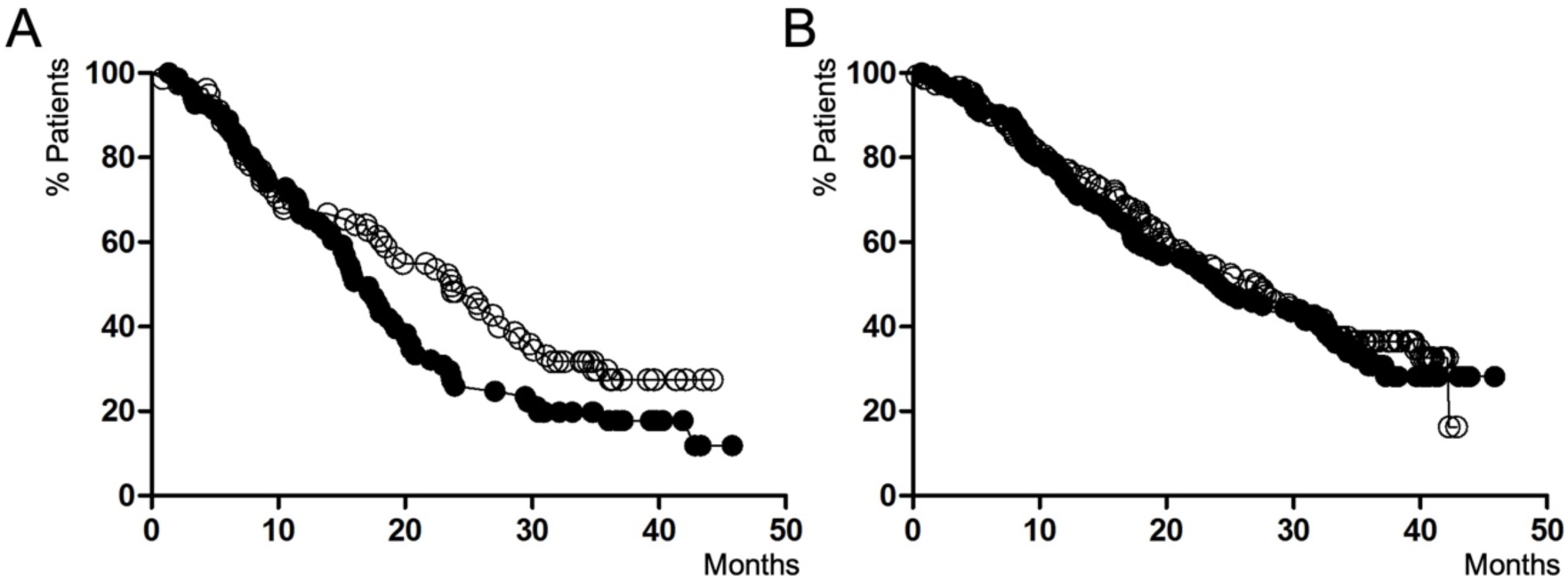
Kaplan–Meier estimate of OS of SPA (A) and SPB (B) after treatment with panitumumab and FOLFOX4 (hollow circles) or FOLFOX4 alone (solid circles).

To explore the parameters contributing to the observed differences in overall survival (OS) between SPA and SPB, we conducted a comparative analysis of clinical and analytical characteristics. The significant differences are summarized in Table 1. A feature contribution analysis identified 13 key distinguishing factors between the subpopulations. Patients in SPA had a higher prevalence of liver metastasis and exhibited elevated levels of ALP, LDH, platelets, WBC, and CEA compared to SPB.

**Table 1.** Significant differences on clinical and laboratory characteristics between SPA* and SPB. All variable values correspond to confidence interval 95%.

| Variable | SPA | SPB | P value |
| --- | --- | --- | --- |
| N | 162 | 298 | - |
| Weight (Kg) | 68.0-72.9 | 73.3-77.6 | .002 |
| BSA (m<sup>2</sup>) | 1.76-1.84 | 1.83-1.88 | .01 |
| BMI | 24.2-25.6 | 25.9-26.9 | <.001 |
| Albumin (g/L) | 3.73-3.84 | 3.88-3.95 | <.001 |
| ALP (UI/L) | 263-313 | 141-164 | <.001 |
| Creatinine (μmol/L) | 0.82-0.86 | 0.86-0.89 | .006 |
| LDH (UI/L) | 795-971 | 340-420 | <.001 |
| Hemoglobin (g/L) | 11.8-12.1 | 12.3-12.5 | <.001 |
| Platelets (10<sup>9</sup>/L) | 330-353 | 269-268 | <.001 |
| WBC (10<sup>9</sup>/L) | 8.07-8.68 | 6.96-7.35 | <.001 |
| CEA (μg/L) | 741-1704 | 42-177 | <.001 |
| Previous surgery (yes; %) | 78.2-89.3 | 90.7-96.1 | <.001 |
| Liver metastasis (yes; %) | 89.8-97.0 | 80.4-88.5 | .002 |

\* SPA: subpopulation A; SPB: subpopulation B; BSA: body surface area; BMI: body mass index; ALP: alkaline phosphatase; LDH: lactate dehydrogenase; WBC: white blood cells; CEA: carcinoembryonic antigen.

In contrast, patients in SPB were characterized by higher weight, BSA, and BMI, underwent more previous surgeries, and had higher levels of albumin, creatinine, and hemoglobin compared to SPA. It is important to highlight that in both subpopulations, levels of ALP, CEA, and LDH were elevated above the normal range, whereas albumin, creatinine, hemoglobin, platelets, and white blood cells remained within normal ranges. This indicated that the system could detect subtle differences between parameters, even when these values fell within normal ranges.

Altogether, these results suggest that the ML framework effectively identified two distinct subpopulations within this cohort, each exhibiting different OS outcomes following treatment with panitumumab/FOLFOX4 or FOLFOX4 alone. In SPA, the addition of panitumumab to FOLFOX4 provided a survival advantage over FOLFOX4 alone, whereas in SPB this survival benefit was not observed. Consequently, we have considered SPA as the subpopulation that benefited from panitumumab treatment, whereas SPB represented the subpopulation that did not experience a survival advantage from the addition of panitumumab to FOLFOX4. Additionally, the ML framework determined that variables such as weight, BSA, BMI, previous surgery, liver metastasis, albumin, ALP, creatinine, LDH, hemoglobin, platelets, WBC, and CEA were associated with these subpopulations, contributing to their differing survival outcomes.

### Generation of PROCC (Panitumumab Response Optimizer for Colorectal Cancer patients) score

With the aim of developing a predictive score capable of identifying patients who would benefit from panitumumab in combination with chemotherapy, that would require fewer input variables than those used for the ML framework and would be easy for physicians to use and understand, we decided to utilize only the baseline values of the differential variables between SPA and SPB detected by ML as input variables in a multivariable logistic regression model. From the thirteen input variables, the independent predictors of panitumumab benefit were identified as ALP, LDH, platelets and CEA (Table 2). The PROCC score was then calculated based on the odds ratio (OR) obtained from the logistic regression analysis.

**Table 2.**
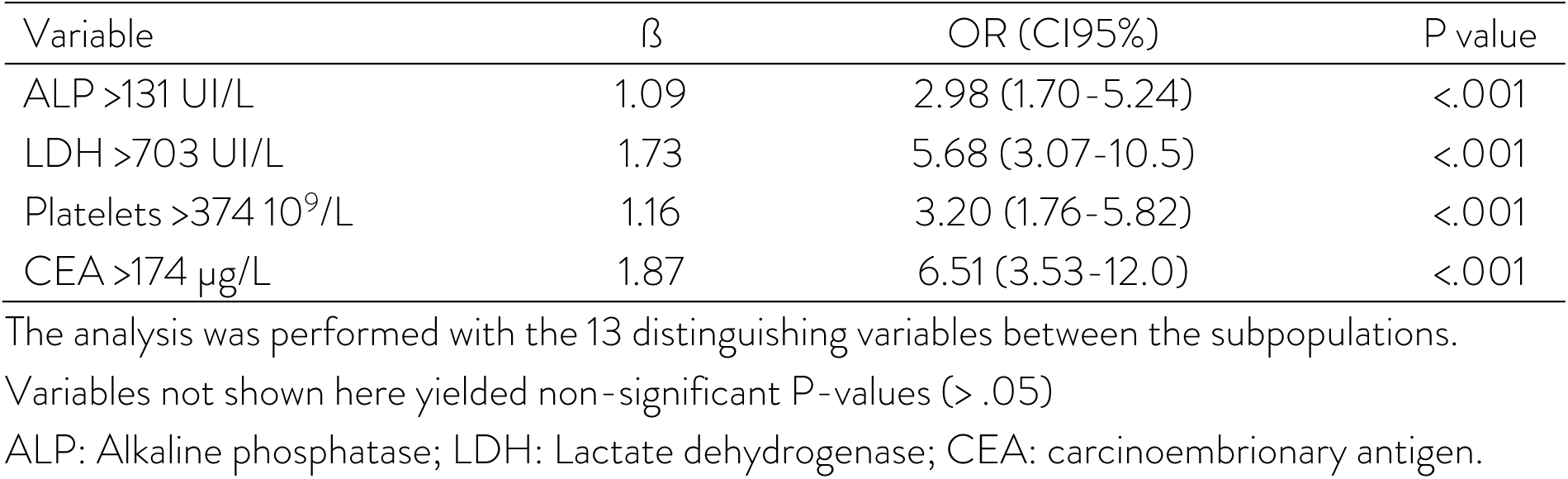
Logistic regression analysis of predictors of panitumumab benefit.

| Variable | $\beta$ | OR (CI95%) | P value |
| --- | --- | --- | --- |
| ALP >131 UI/L | 1.09 | 2.98 (1.70-5.24) | <.001 |
| LDH >703 UI/L | 1.73 | 5.68 (3.07-10.5) | <.001 |
| Platelets >374 $10^9/L$ | 1.16 | 3.20 (1.76-5.82) | <.001 |
| CEA >174 $\mu g/L$ | 1.87 | 6.51 (3.53-12.0) | <.001 |

The analysis was performed with the 13 distinguishing variables between the subpopulations.
Variables not shown here yielded non-significant P-values (> .05)
ALP: Alkaline phosphatase; LDH: Lactate dehydrogenase; CEA: carcinoembryonary antigen.

To enhance clarity and facilitate practical application, each OR was rounded to the nearest integer or 0.5, as appropriate. The resulting PROCC score ranged from 0 to 18 and was classified as following: 6.5 points for CEA >174 ng/ml, 5.5 for LDH >703 IU/L, 3 points for each APL >131 IU/L and platelets >374 10^9^/L (Table 3). The cut-off for grouping the population according to panitumumab benefit or not (binary classification) were manually determined, considering the positivity rates for panitumumab benefit within each category. For instance, only 3 of the 150 patients with 0 points were classified as belonging to the panitumumab benefited group while 19 out of 20 with 18 points were classified to the panitumumab benefited group.

**Table 3.**
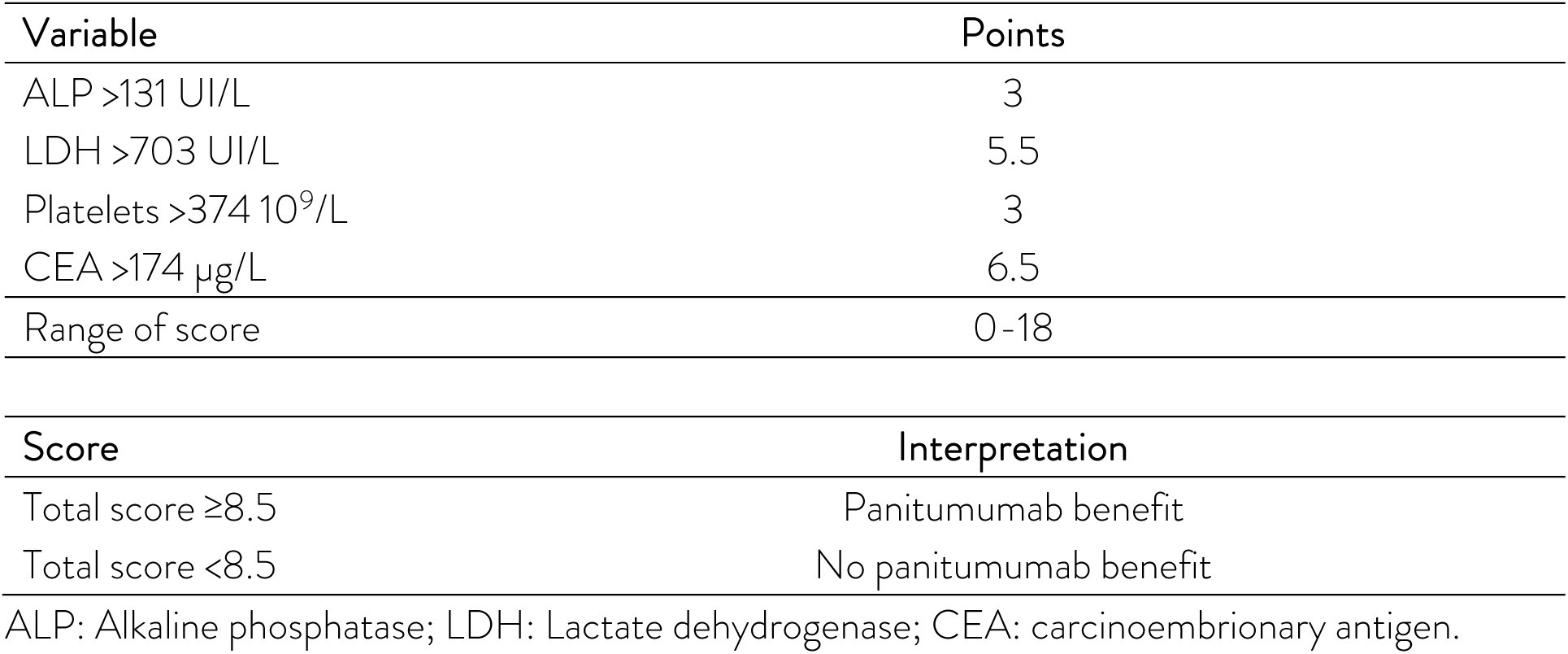
Proposed scoring system and clinical interpretation.

| Variable | Points |
| --- | --- |
| ALP $>131$ UI/L | 3 |
| LDH $>703$ UI/L | 5.5 |
| Platelets $>374 \times 10^9/L$ | 3 |
| CEA $>174 \mu g/L$ | 6.5 |
| Range of score | 0-18 |

| Score | Interpretation |
| --- | --- |
| Total score $\geq 8.5$ | Panitumumab benefit |
| Total score $< 8.5$ | No panitumumab benefit |

ALP: Alkaline phosphatase; LDH: Lactate dehydrogenase; CEA: carcinoembryonary antigen.

When TRDS was classified by PROCC to predict panitumumab benefit, the area under the ROC curve was 0.81 (95% CI: 0.77 to 0.85) (Figure 3). The optimal cut-off for stratifying patients by their likelihood of benefiting from panitumumab was determined using Youden’s index and accuracy metrics, complemented by a manual review of positivity rates across score categories. A threshold of ≥8.5 was identified, yielding a sensitivity of 67.1%, a specificity of 88.0%, and an overall accuracy of 80.8% (Youden’s index = 0.55). This cut-off was subsequently applied to the TRDS cohort for further validation and subgroup analysis.

**Figure 3.**
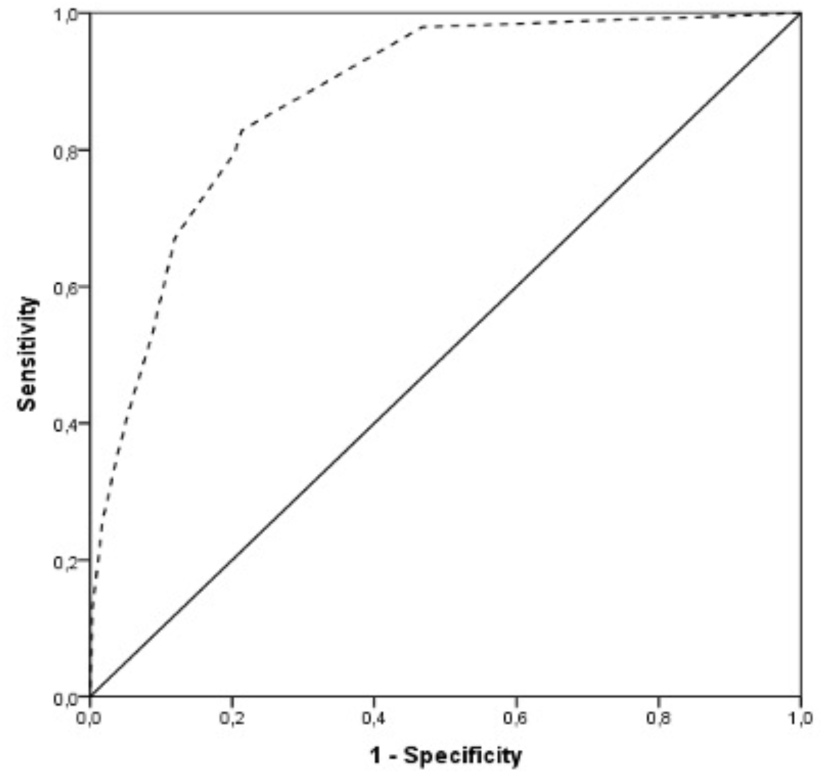
Receiver-Operating Characteristic Curve (ROC) of PROCC scoring system for predicting survival benefit of adding panitumumab to chemotherapy treatment.

### Application of PROCC to the TRDS cohort

To confirm that PROCC could identify patients who benefit from receiving panitumumab, the score was applied to TRDS patients. Out of the total population, 143 had PROCC score values ≥8.5, while 317 had values <8.5. In patients with score values ≥8.5, 76 patients received panitumumab/FOLFOX4, while 67 were treated with FOLFOX4 alone. In patients with score values <8.5, 156 patients were treated with panitumumab/FOLFOX4 and 161 with FOLFOX4 alone. Subsequently, survival analyses were conducted for both subpopulations based on treatment with panitumumab/FOLFOX4 or FOLFOX4 alone. As illustrated in Figure 4A and Figure 4C, patients with PROCC ≥8.5 treated with panitumumab/FOLFOX4 exhibited a significantly reduced risk of progression (HR 0.67; 95% CI 0.47-0.97; p=.03) and death (HR 0.65; 95% CI 0.43-0.98; p=.04), with a median PFS of 9.3 months (95% CI 6.0-9.8) and OS of 23.6 months (95% CI 15.2-32), compared to those receiving FOLFOX4 alone. This latter group showed a median PFS of 6.7 months (95% CI 5.4-9.1) and OS of 15.9 months (95% CI 12.9-19.0).

**Figure 4.**
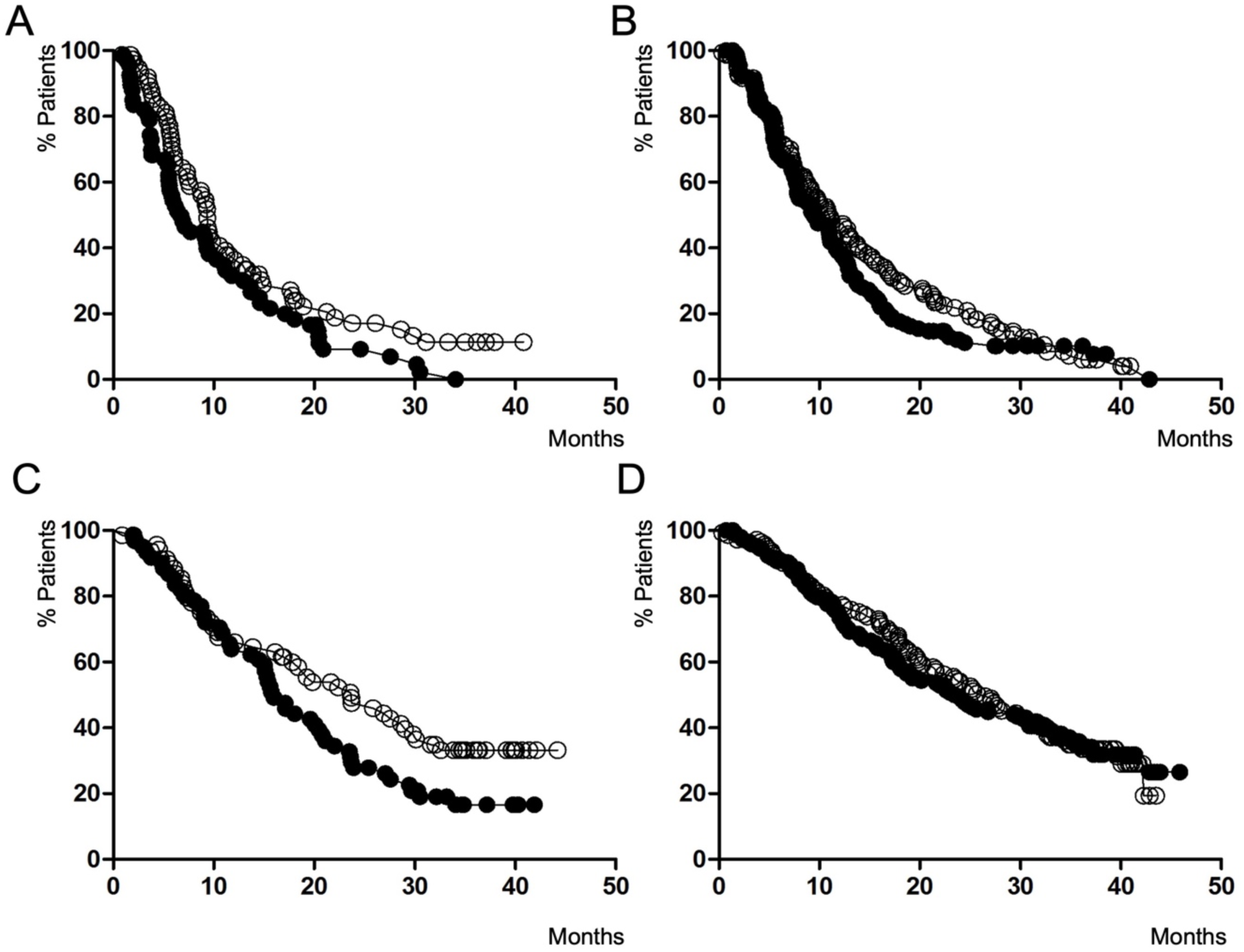
Kaplan–Meier estimate of PFS and OS of patients of the training dataset according to PROCC after treatment with panitumumab and FOLFOX4 (hollow circles) or FOLFOX4 alone (solid circles). (A) PFS for patients with PROCC ≥ 8.5 (B) PFS for patients with PROCC < 8.5 (C) OS for patients with PROCC ≥ 8.5 (D) OS for patients with PROCC < 8.5.

In contrast, patients with PROCC <8.5 showed no significant differences in PFS (HR 0.84; 95% CI 0.65-1.07; p=.1) or OS (HR 0.97; 95% CI 0.72-1.30; p=.8) between the two treatments (Figures 4B and 4D). For those treated with panitumumab/FOLFOX4, the median PFS was 11.0 months (95% CI 7.6-11.2), and the OS was 26.4 months (95% CI 21.8-31.1). For the FOLFOX4 group, the median PFS was 9.4 months (95% CI 7.3-10.7), and the OS was 23.9 months (95% CI 17.9-30.0). These results demonstrated that PROCC was able to identify patients for whom the addition of panitumumab to chemotherapy was beneficial in terms of clinical outcomes.

### Validation of PROCC on the VALDS cohort

To validate PROCC in an independent dataset that was not utilized during its development, we applied the score to a cohort of 479 patients from a new clinical study (NCT00339183; see Materials and Methods). Baseline characteristics of this cohort are detailed in Supplementary Table 2 and Supplementary Figure 3. Among the total population, 141 patients had PROCC scores ≥8.5, while 338 patients had scores <8.5. In the group with scores ≥8.5, 75 patients received panitumumab/FOLFIRI, and 66 patients were treated with FOLFIRI alone. In the group with scores <8.5, 167 patients were treated with panitumumab/FOLFIRI, while 171 patients received FOLFIRI alone. Subsequent survival analyses were conducted based on treatment with panitumumab/FOLFIRI or FOLFIRI alone in both subpopulations. As shown in Figures 5A and 5C, patients with PROCC scores ≥8.5 treated with panitumumab/FOLFIRI demonstrated a significantly reduced risk of progression (HR 0.48; 95% CI 0.33-0.70; p=.0002) and death (HR 0.60; 95% CI 0.42-0.87; p=.007), with a median PFS of 5.9 months (95% CI 4.47-7.49) and OS of 13.6 months (95% CI 9.40-16.5), compared to those receiving FOLFIRI alone. This latter group showed a median PFS of 3.6 months (95% CI 2.13-3.78) and OS of 7.3 months (95% CI 5.70-8.20). Conversely, patients with PROCC <8.5 showed no significant differences in PFS (HR 0.88; 95% CI 0.70-1.10; p=.2) or OS (HR 0.93; 95% CI 0.73-1.17; p=.5) between the two treatments (Figures 5B and 5D). In this group, patients treated with panitumumab/FOLFIRI had a median PFS of 7.3 months (95% CI 5.58-7.49) and OS of 15.5 months (95% CI 11.8-18.1), while those receiving FOLFIRI alone had a median PFS of 5.32 months (95% CI 3.81-5.68) and OS of 14.4 months (95% CI 11.3-16.6). These findings further demonstrated that PROCC could effectively identify patients who benefit from the addition of panitumumab to chemotherapy in terms of improved clinical outcomes in a new clinical dataset.

**Figure 5.**
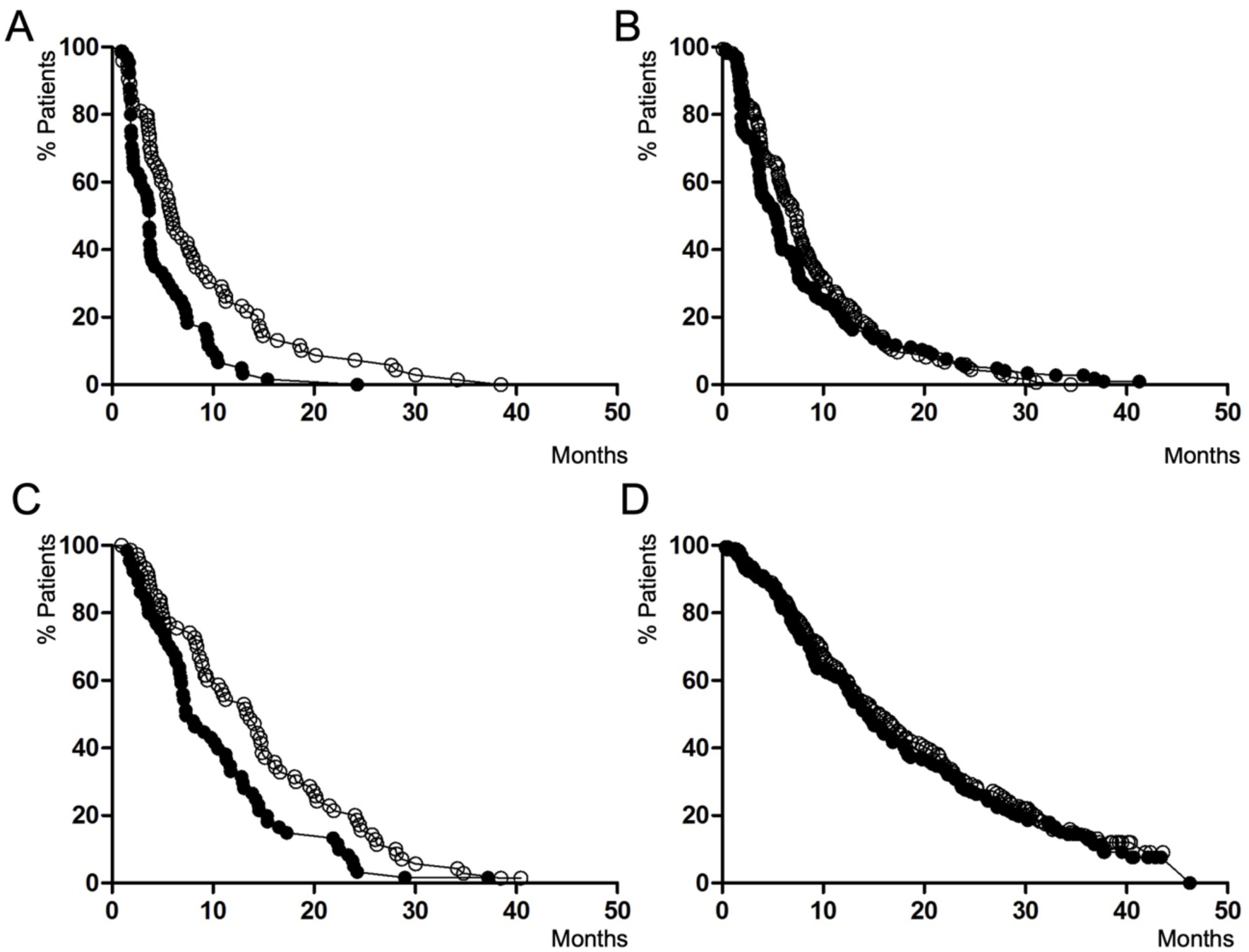
Kaplan–Meier estimate of PFS and OS of patients of the test dataset according to PROCC after treatment with FOLFIRI and panitumumab (hollow circles) or FOLFIRI alone (solid circles). (A) PFS for patients with PROCC ≥ 8.5 (B) PFS for patients with PROCC < 8.5 (C) OS for patients with PROCC ≥ 8.5 (D) OS for patients with PROCC < 8.5.

## DISCUSSION

In this study, we have developed PROCC, a predictive score using a disease mechanism-driven and data-based approach to identify KRAS WT mCRC patients who will significantly benefit from adding panitumumab to chemotherapy (FOLFOX or FOLFIRI) as first- or second-line therapy. Our analysis shows that panitumumab-related benefits are primarily observed in patients with high PROCC scores, while those with low scores do not experience such improvements. PROCC effectively identifies a subpopulation that generally has poorer survival outcomes with chemotherapy alone but sees markedly improved results with the addition of panitumumab.

Notably, PROCC only requires pre-treatment values of four commonly measured blood metrics in mCRC patients: CEA, LDH, ALP, and platelets, making it a simple yet powerful tool for optimizing panitumumab treatment strategies. These four variables were identified through the combined use of ML techniques and classical biostatistics. The 4-predictor-based scoring system demonstrated strong discrimination. Internal and external validation confirmed the model’s reliability, further supporting its use as a potential effective tool for clinical decision-making in this patient population. PROCC score is aligned with a precision oncology approach, that would aim to maximizing efficacy while minimizing unnecessary side effects.

In current practice, panitumumab administration for the treatment of mCRC relies heavily on KRAS status and tumor sidedness to predict patient benefit on this therapy^16, 21^. Different studies have highlighted the critical role of KRAS mutation status in determining which patients with mCRC will benefit from addition of panitumumab to chemotherapy in first- and second line treatment. The PRIME phase 3 trial showed that patients with KRAS WT tumors experienced longer PFS and OS when treated with panitumumab plus FOLFOX4, compared to FOLFOX4 alone^11, 17^. These findings were confirmed in second-line settings, where panitumumab plus FOLFIRI also significantly improved PFS in KRAS WT mCRC patients^12^. Additional studies demonstrated significant differences in panitumumab’s efficacy based on tumor sidedness, with left-sided KRAS WT tumors showing better responses to panitumumab combined with chemotherapy (FOLFOX or FOLFIRI) compared to right-sided tumors^19, 20^. This is likely due to a higher prevalence of molecular alterations in right-sided tumors that confer resistance to anti-EGFR therapies. As a result, panitumumab is primarily recommended for left-sided tumors, while alternative therapies like bevacizumab are preferred for right-sided mCRC due to more consistent efficacy in this subgroup^22^. Despite its effectiveness, combinations of panitumumab and chemotherapy are associated with serious adverse effects, including skin toxicity, diarrhea, neutropenia, and hypomagnesemia, which can lead to treatment discontinuation of 23% for patients treated with panitumumab/FOLFOX and 29% for those treated with panitumumab/FOLFIRI^19, 23, 24^. These findings emphasize the need for improved patient selection to maximize benefits and minimize toxicity, suggesting that factors beyond tumor sidedness and mutational status should be explored to better identify patients who will truly benefit from panitumumab treatment.

The primary objective in developing PROCC was to create a reliable predictive score for identifying KRAS WT mCRC patients most likely to benefit from panitumumab, supporting clinicians in making more informed treatment decisions regarding the inclusion of panitumumab with chemotherapy. PROCC consistently demonstrated, across two different datasets, that patients with scores ≥8.5 had improved survival outcomes (PFS and OS) when treated with panitumumab plus chemotherapy compared to those treated with chemotherapy alone, while those with scores <8.5 did not derive additional benefits from panitumumab addition, suggesting that chemotherapy alone or alternative treatments may be a more appropriate option. Indeed, these latter patients tend to have a relatively favorable prognosis, and panitumumab addition achieve survival results not different from the outcome of patients receiving chemotherapy alone. For this subgroup, adding panitumumab would not yield further survival benefits but could lead to unnecessary adverse effects and treatment costs. Our findings, thus, suggest that all panitumumab benefits observed in the two studied cohorts are confined to patients with PROCC scores ≥8.5. This patient population represented about 30% of KRAS WT mCRC patients (TRDS= 143/460 patients; VALDS= 141/479 patients). These patients are the only likely to benefit from panitumumab, while the remaining 70% could potentially avoid unnecessary panitumumab side effects if PROCC-based treatment decisions were applied. Thus, PROCC would allow a more selective and efficient use of panitumumab. Furthermore, PROCC shows robust predictive ability across different clinical contexts, being developed in a first-line setting (PRIME trial), and validated in a second-line phase 3 trial. Thus, this novel, data-driven approach can complement traditional biomarkers like KRAS status or tumor sidedness and could improve treatment strategies as it can help to identify KRAS WT mCRC patients likely to benefit from panitumumab in different clinical settings.

PROCC was developed using a sequential approach, starting with an unsupervised machine learning model to identify clinical characteristics associated with poor clinical outcome, followed by a logistic regression to generate the predictive score to identify KRAS WT mCRC patients likely to benefit from adding panitumumab to chemotherapy. This strategy allowed us to use a “black-box” system to identify clinical and analytical features associated with a particular poor clinical outcome and subsequently provide “explainability” to the result with a “crystal-clear box”, through the generation of a classic predictive score, a system that healthcare professionals are accustomed to using. In this way, we aim to reduce the uncertainty generated when working with complex systems in which it is challenging to understand the results. The use of unsupervised ML enabled us to identify a subpopulation of patients (SPA) with certain characteristics associated with poor clinical outcome. These patients, representing approximately 30% of the KRAS WT population, showed a higher prevalence of liver metastases and had elevated levels of ALP, LDH, CEA, platelets, and WBC compared to the other patients (SPB). Notably, the system was able to detect differences between both subpopulations for variables in abnormal ranges (ALP, LDH, or CEA) and even in variables within normal ranges (platelets and WBC), differences that are impossible to identify through clinical criteria. For instance, 95% CI for WBC in SPA was 8.07-8.68 10^9^/L compared to 6.96-7.35 10^9^/L in SPB (p<.001). Interestingly, these clinical differences were associated with improved overall survival when panitumumab was administered alongside chemotherapy. These results suggest that varied responses to panitumumab treatment are influenced by clinical factors not captured by traditional inclusion criteria in clinical trials. This highlights the model’s ability to detect intrinsic features that distinguish responsive from non-responsive patients.

The thirteen differentiating variables between SPA and SPB were used to generate a predictive score for response to panitumumab. Our logistic regression analysis identified that four out of thirteen are statistically significant predictors: ALP, LDH, CEA and platelets. The identified predictors in this study align with the findings of previous research, particularly regarding CEA. In our study, having pretreatment levels of CEA >174 µg/L demonstrated the strongest association among these predictors, with an adjusted OR of 6.5 (95% CI, 3.53-12.0; p<.001). The correlation between CEA levels and the effectiveness of panitumumab in KRAS WT mCRC patient is not well studied in the scientific literature. In these patients, high levels of CEA have been correlated with a higher tumor burden and with more aggressive disease and worse prognosis, which is in line with the findings of our current study^25^. However, the correlation between CEA levels and the effectiveness of panitumumab is more nuanced as apparently pretreatment CEA levels alone do not directly predict the efficacy of the drug^26^. Interestingly, our study also identified LDH, ALP and platelets as new predictors for panitumumab benefit. To our knowledge, there is no direct correlation reported between LDH and ALP levels and response to panitumumab. There is only one phase II study reporting that LDH levels may predict early responses to panitumumab combined with fluorouracil treatment in KRAS WT mCRC patients^27^. Nonetheless, these two predictors are biologically plausible, as they are frequently linked with more aggressive tumor behavior, increased tumor burden, and poorer prognosis in many cancers, including mCRC^28, 29^. In reference to platelets, it is well known that they play a pivotal role in cancer progression with a decrease in overall survival and poorer prognosis of mCRC patients^30, 31^. However, the precise mechanism by which elevated levels of these three markers are associated with improved survival outcomes from panitumumab treatment remains unclear, therefore, additional research is warranted to assess these correlations.

Several limitations should be noted in this study. First, the development of PROCC was based on data from a single retrospective clinical trial involving 460 patients of the total 1,183 patients. Although this sample size was relatively modest, it provided sufficient statistical power to identify significant variables within the cohort. The use of machine learning to pre-select variables for inclusion in the multivariable logistic regression greatly facilitated the identification of key factors, and this strategy has been successfully applied in other cancer patient cohorts, including prostate and lung cancer ^32, 33^. Second, the external validation of PROCC was limited to a single dataset of 479 patients, and further research is needed to validate this score in other clinical settings and datasets, involving panitumumab or related anti-EGFR therapies (e.g., cetuximab, necitumumab, etc.). Third, the score relied on a complete-case analysis without data imputation, meaning that in cases where one or more of the four variables were missing at baseline, the score was calculated based on the remaining values. This could have led to some patients being incorrectly classified as having a score of less than 8 when they should have been considered high scorers. However, this affected only a small fraction (2.2%) of the TRDS cohort and did not impact the validation set, which had no missing data. Lastly, the relationship between PROCC and tumor sidedness could not be studied due to the lack of data on tumor location. Despite these limitations, the study has several important strengths. To the best of our knowledge, no prior study has established a prediction model for assessing the benefit of adding panitumumab to chemotherapy in patients with KRAS WT mCRC. Therefore, our work represents an early attempt to develop and validate a clinical score that correlates panitumumab efficacy with survival outcomes based on clinical data. Moreover, PROCC incorporates variables commonly measured in routine clinical practice, which enhances its generalizability and potential applicability in everyday clinical settings.

In conclusion, based on pretreatment levels of CEA, ALP, LDH, and platelet counts, the PROCC predictive score may serve as a valuable tool for identifying KRAS WT mCRC patients likely to benefit from adding panitumumab to chemotherapy in either first or second-line settings. Given the widespread use of panitumumab regimens, PROCC has the potential to significantly impact treatment decisions by improving efficacy and minimizing side effects. This four-predictor-based scoring system demonstrated strong discrimination and calibration performance, helping prioritize patients most likely to gain from panitumumab treatment. Moreover, the ease of measuring these four markers makes PROCC suitable for use in real-world environments, as the tests are inexpensive, noninvasive, and convenient. Additional prospective validation in various clinical settings is necessary to confirm its broader applicability.

## Data Availability

Datasets were obtained from www.projectdatasphere.org

## ACKNOWLEDGEMENTS

We are grateful to www.projectdatasphere.com that allow us to produce this research through access to the high-quality datasets used in the study.

## SUPPLEMENTARY MATERIAL

**Supplementary Table 1.** Clinical characteristics of patients on the training dataset (TRDS)

| Variable |  |  |
| --- | --- | --- |
| N | 460 |  |
|  | Mean | CI95% |
| Age (years) | 60.7 | 59.7-61.7 |
| Weight (Kg) | 73.4 | 72.0-74.8 |
| Height (cm) | 167 | 167-168 |
| BSA* (m<sup>2</sup>) | 1.83 | 1.81-1.86 |
| BMI | 25.9 | 25.5-26.3 |
| Albumin (g/L) | 3.93 | 3.88-3.98 |
| ALP (UI/L) | 204 | 184-223 |
| Creatinine (μmol/L) | 0.88 | 0.86-0.89 |
| LDH (UI/L) | 648 | 573-724 |
| Hemoglobin (g/L) | 12.4 | 12.3-12.5 |
| Platelets (10<sup>9</sup>/L) | 320 | 309-331 |
| WBC; 10<sup>9</sup>/L | 8.33 | 8.04-8.62 |
| CEA (μg/L) | 496 | 317-675 |
| PFS | 9.38 | 7.65-9.48 |
| OS | 22.9 | 18.8-23.6 |
|  | N | % |
| Sex |  |  |
| Female | 163 | 35.4 |
| Male | 297 | 64.6 |
| ECOG 0/1 | 434 | 94.3 |
| Treatment |  |  |
| P/FOL | 232 | 50.4 |
| FOL | 228 | 49.6 |
| Previous surgery (yes) | 417 | 90.6 |
| Liver metastasis | 406 | 88.2 |

\* BSA: body surface area; BMI: body mass index; ALP: alkaline phosphatase; LDH: lactate dehydrogenase; WBC: white blood cells; CEA: carcinoembryonary antigen; PFS: progression-free survival; OS: overall survival; P/FOL: panitumumab/FOLFOX4 combination; FOL: FOLFOX4.

**Supplementary Table 2.** Clinical and analytical characteristics of patients on the validation dataset (VALDS)

| Variable |  |  |
| --- | --- | --- |
| N | 479 |  |
|  | Mean | CI95% |
| Age (years) | 60.5 | 59.6-61.4 |
| ALP* (UI/L) | 249 | 234-264 |
| LDH (UI/L) | 491 | 460-522 |
| Platelets (10<sup>9</sup>/L) | 261 | 251-270 |
| CEA (µg/L) | 403 | 198-607 |
| PFS | 5.58 | 4.53-5.78 |
| OS | 13.3 | 11.2-13.8 |
|  | N | % |
| Sex |  |  |
| Female | 179 | 37.3 |
| Male | 300 | 63.6 |
| ECOG 0/1 | 451 | 94.1 |
| Treatment |  |  |
| P/FOLFIRI | 242 | 50.5 |
| FOLFIRI | 237 | 49.4 |

\* ALP: alkaline phosphatase; LDH: lactate dehydrogenase; CEA: carcinoembryonary antigen; PFS: progression-free survival; OS: overall survival; P/FOLFIRI: panitumumab/FOLFIRI combination.

**Supplementary Figure 1.**
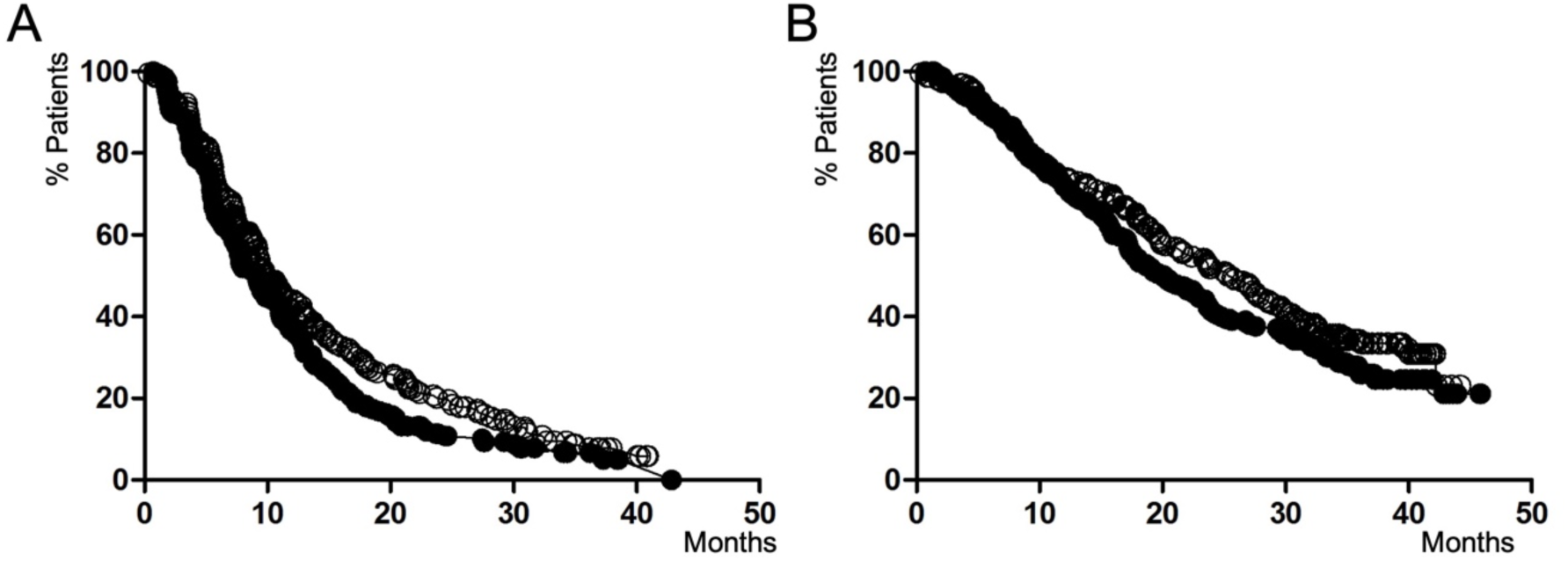
Kaplan-Meier estimates of PFS (A) and OS (B) of patients on the training set after treatment with FOLFOX4 and panitumumab (hollow circles) or FOLFOX4 alone (solid circles). (A) Median PFS for the panitumumab/FOLFOX4 arm was 9.9 months (95%CI 7.9-10.6). Median PFS for the FOLFOX4 arm was 6.7 months (95%CI 7.9-9.3). Patients treated with panitumumab/FOLFOX4 had a lower risk of progression compared to those treated with FOLFOX4 alone (HR 0.79 95%CI 0.64-0.97; p=.02). (B) Median OS for the panitumumab/FOLFOX4 arm was 25.6 months (95%CI 19.4-27.0). Median OS for the FOLFOX4 arm was 20.1 months (95%CI 17.1-22.5). No significant differences on OS between panitumumab/FOLFOX4 and FOLFOX4 arms were observed (HR: 0.83 95%CI 0.66-1.04; p=.1).

**Supplementary Figure 2.**
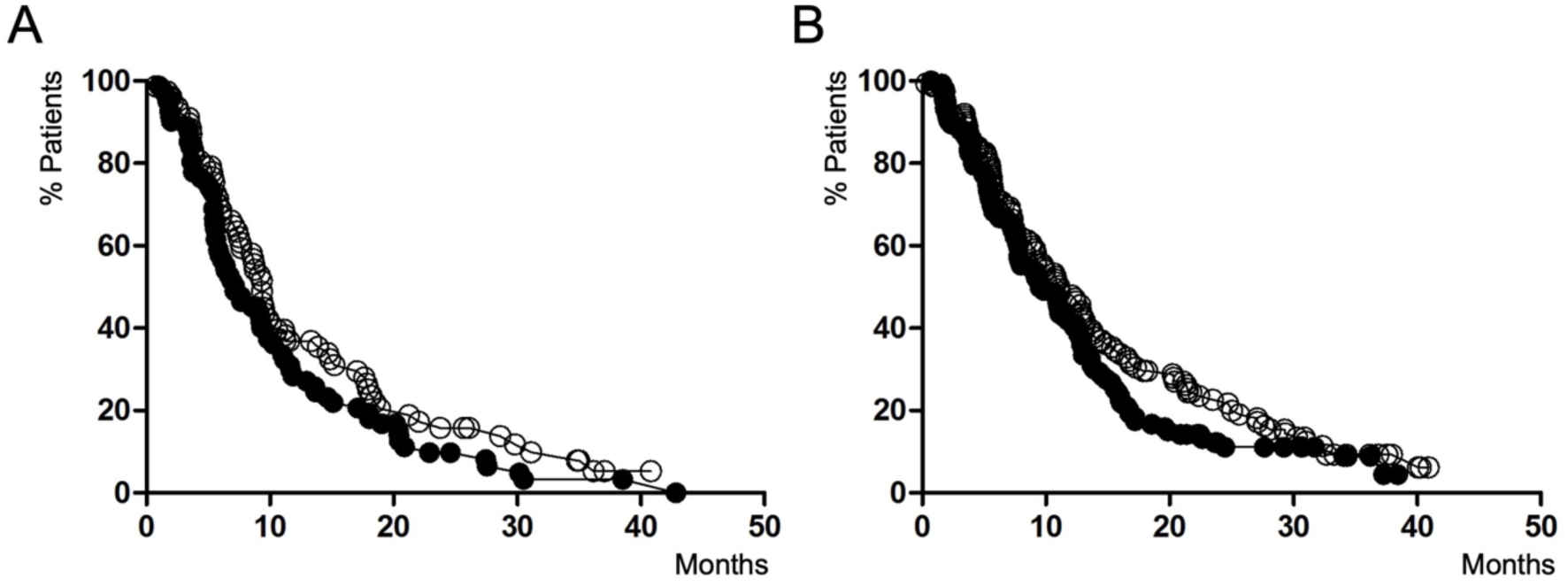
Kaplan-Meier estimate of PFS for SPA (A) and SPB (B) after treatment with panitumumab and FOLFOX4 (hollow circles) or FOLFOX4 alone (solid circles). (A) Median PFS for the panitumumab/FOLFOX4 arm was 9.3 months (95%CI 7.0-10.0). Median PFS for the FOLFOX4 arm was 7.1 months (95%CI 5.5-9.2). No significant differences on PFS between panitumumab/FOLFOX4 and FOLFOX4 arms were observed (HR: 0.77 95%CI 0.55-1.08; p=.1). (B) Median PFS for the panitumumab/FOLFOX4 arm was 11.0 months (95%CI 7.3-12.0). Median PFS for the FOLFOX4 arm was 9.1 months (95%CI 7.2-10.8). No significant differences on PFS between panitumumab/FOLFOX4 and FOLFOX4 arms were observed (HR: 0.80 95%CI 0.62-1.04; p=.1).

**Supplementary Figure 3.**
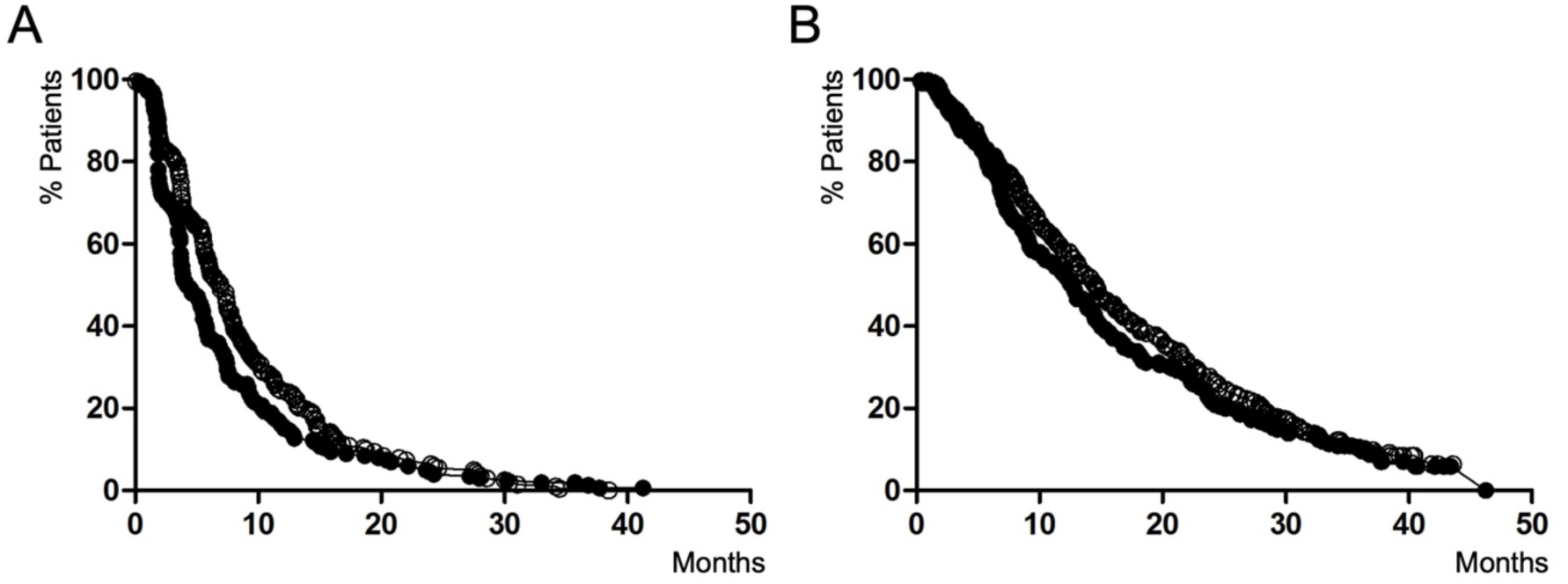
Kaplan-Meier estimate of PFS (A) and OS (B) of patients on the validation set after treatment with panitumumab and FOLFIRI (hollow circles) or FOLFIRI alone (solid circles). (A) Median PFS for the panitumumab/FOLFIRI arm was 6.9 months (95%CI 5.5-7.3). Median PFS for the FOLFIRI arm was 4.2 months (95%CI 3.6-5.2). Patients treated with panitumumab/FOLFIRI had a lower risk of progression compared to those treated with FOLFIRI (HR 0.77 95%CI 0.64-0.93; p=.009). (B) Median OS for the panitumumab/FOLFIRI arm was 14.5 months (95%CI 11.6-15.1). Median OS for the FOLFIRI arm was 12.6 months (95%CI 9.1-13.0). No significant differences on OS between panitumumab/FOLFIRI and FOLFIRI groups were observed (HR: 0.88 95%CI 0.72-1.07; p=.2).

